# Exploring impacts of long-COVID-19 on the lungs: A triad of PET scans for perfusion, inflammation and tissue remodeling

**DOI:** 10.1101/2025.06.13.25328911

**Authors:** Olivia Wegrzyniak, Olof Eriksson, Emil Ekbom, Robert Frithiof, Michael Hultström, Mark Lubberink, Andrei Malinovchi, Jonathan Sigfridsson, Irina Velikyan, Viola Wilson, Gunnar Antoni, Miklos Lipcsey

## Abstract

**Background:** COVID-19 patients may experience long-lasting complications, particularly impairment of lung function. The persistence of inflammation may lead to tissue remodeling, but the impact on the lungs in these patients remains unclear.

**Research Question:** Is there evidence of neutrophil-mediated inflammation, tissue remodeling, and altered lung perfusion in individuals with long COVID-19 experiencing dyspnea and reduced diffusion capacity? Additionally, can the PET imaging findings be correlated with proteomic data reflecting inflammatory status and clinical information?

**Study Design and Methods:** Patient experiencing persistent dyspnea and reduced diffusion capacity for carbon monoxide 20 to 30 months after severe COVID-19 illness were recruited and underwent sequential PET scans using [^15^O]water, [^68^Ga]Ga-FAPI-46, and [^11^C]NES. Clinical lung function and 6-minute walking tests were performed before the scans. Blood samples were collected for plasma analysis using proximity extension analysis, which enabled the measurement of 96 inflammation-related biomarkers.

**Results:** Six male patients were recruited (age 64 ± 7 [median ± range] years). Patients with known impaired lung function prior to COVID-19 illness were excluded. [^15^O]water did not show a clear reduction in areas with signs of inflammation or tissue remodeling. However, all patients showed uptake of [^68^Ga]Ga-FAPI-46 and [^11^C]NES in the lungs, especially in areas associated with inflammation and remodeling. Patients were categorized into two groups based on tracer uptake: a low-uptake group and a high-uptake group. While the plasma inflammatory profiles differed between long-COVID-19 patients and healthy individuals, there was no clear correlation between the lung uptake of [^68^Ga]Ga-FAPI-46 and [^11^C]NES in long-COVID-19 patients. Similar observations were made regarding clinical parameters.

**Interpretation:** Sequential PET scans using [^15^O]water, [^68^Ga]Ga-FAPI-46, and [^11^C]NES, allowed observation of signs of neutrophil-mediated inflammation and tissue remodeling in patients with long-COVID-19. However, no association with lung perfusion was demonstrated. Additionally, the results from the PET scans could not be clearly associated with clinical parameters or inflammatory proteomics.

## Introduction

Coronavirus disease 2019 (COVID-19), caused by severe acute respiratory syndrome coronavirus 2 (SARS-CoV-2), is a highly transmissible virus that emerged in late 2019, leading to a global pandemic with widespread health, economic, and societal impacts^1,2^. Patients diagnosed with COVID-19 often experience long-term complications, including prolonged impairment of lung function, especially after severe illness^3–5^.

Neutrophils play a major role in the immune response to COVID-19 infection ^6,7^, and may also play a role in long-COVID-19^8^. Positron emission tomography/computed tomography (PET/CT) using [^11^C]NES, which targets neutrophil elastase (NE) secreted by neutrophils, in patients with severe COVID-19 revealed high uptake of [^11^C]NES in the lungs^9^. NETosis, which involves the formation of neutrophil extracellular traps (NETs), contribute to the development of complications in lung diseases.^10,11^ Persistent inflammation post-COVID-19, particularly with active NETosis, may damage the alveolar epithelium and endothelium, potentially resulting in fibrosis.

Fibrosis is characterized by the accumulation of persistently activated myofibroblasts, which are responsible for extracellular matrix deposition and remodeling of the affected tissue. The endopeptidase fibroblast activation protein (FAP) is expressed at the surface of such cells. PET imaging with the FAP inhibitor [^68^Ga]Ga-FAPI-04 showed tracer accumulation in fibrotic areas of the lungs in patients with interstitial lung diseases (ILDs)^12^.

The inflammatory and fibrogenesis processes in long-COVID-19 remain unclear. In addition, while inflammation can be treated, pulmonary fibrosis is primarily preventable but fibrotic tissue cannot be removed and no effective treatment exists. Early detection of ongoing fibrosis or tissue remodeling is crucial and allows for preventive actions to be taken before irreversible damage to the lungs occurs. Therefore, there is a need for a non-invasive technique to study long-COVID-19 long-term lung function impairment and improve patient stratification for targeted therapeutic interventions. We hypothesized that neutrophil-mediated inflammation as well as tissue remodeling were present in the lungs of patients with dyspnea and decreased diffusion capacity after long-COVID-19.

Here, we present the results of a single center, pilot study examining the potential of [^11^C]NES and [^68^Ga]Ga-FAPI-46 PET-CT scans in identifying patients with persistent inflammation or early fibrosis. Additionally, we evaluated lung perfusion using [^15^O]water PET scans to identify potential correlations with patients’ clinical symptoms and the respective uptake observed in [^68^Ga]FAPI-46 and [^11^C]NES imaging. Venous blood samples were taken for proximity extension assay analysis, for correlation of PET- and patient clinical data with inflammation biomarkers.

## Material and methods

### Subjects

Approval was obtained from the Swedish Ethical Review Authority (Dnr.: 2021-03218). This study was conducted in accordance with the principles outlined in the Declaration of Helsinki. Patient experiencing persistent dyspnea and reduced diffusion capacity for carbon monoxide 20 to 30 months after severe COVID-19 illness were recruited. The exclusion criterion was : impaired lung function prior to COVID-19 illness. Patient characteristics are presented in Table 1. All participants provided verbal and written informed consent.

**Table 1.** Demographics and clinical characteristics of the long-COVID-19 patients.

| Variable |  |  |
| --- | --- | --- |
| Age (years)* |  | 64 (63.75; 69.25) |
| Sex (n (%)) | Male | 6 (100%) |
| BMI (kg/m<sup>2</sup>)* |  | 30 (26.50; 32.25) |
| Smoking status (n (%)) | Current smoker | 1 (17%) |
|  | Former smoker | 1 (17%) |
|  | Never smoked | 4 (66%) |
| Co-morbid diseases (n (%)) | Hypertension | 6 (100%) |
|  | Diabetes mellitus | 2 (33%) |
|  | Cancer | 0 (0%) |
|  | Asthma | 3 (50%) |
|  | COPD | 0 (0%) |
|  | ILD | 0 (0%) |
|  | Autoimmune Diseases | 0 (0%) |
|  | Chronic kidney failure | 2 (33%) |
|  | Cirrhosis | 0 (0%) |
| Invasive ventilation | IV (n (%)) | 5 (83%) |
|  | Duration IV (n=5) (days)* | 9 (6.50; 26.50) |
| Treatment (n (%)) | Corticosteroid p.o. | 0 (0%) |
|  | Corticosteroid inh. | 5 (83%) |
|  | NSAID | 1 (17%) |
| LoS ICU (days)* |  | 16.50 (14.25; 31) |
| LoS Hospital (days)* |  | 42.50 (27.25; 109) |
| 6MWD (m)* |  | 178 (167; 189.5) |
| Pulmonary function test * | FVC (%) | 88.50 (74.50; 95.50) |
|  | FVC (L) | 3.90 (3.54; 4.28) |
|  | FEV1 (%) | 91.00 (84.00; 101,8) |
|  | FEV1 (L) | 3.21 (2.67; 3.46) |
|  | FEV1/FVC (%) | 81.51 (77.15; 83.45) |
|  | RV (%) | 88.5 (73.50; 102.30) |
|  | RV (L) | 1.82 (1.57; 2.16) |
|  | TLC (%) | 82.50 (78.50; 95.00) |
|  | TLC (L) | 5.92 (5.14; 6.70) |
|  | DLCO (%) | 67 (60.50; 70.75) |

\*Data are presented as median (IQR25; IQR75). BMI, body mass index; COPD, chronic obstructive pulmonary disease; DLCO, diffusing capacity for carbon monoxide; FEV1, forced expiratory volume in 1 s; FEV1/FVC, Tiffeneau-Pinelli index; FVC, forced vital capacity; ICU, intensive care unit; inh, inhaled; ILD, Interstitial lung disease; IV, invasive ventilation; LoS, length of stay; NSAID, nonsteroidal anti-inflammatory drugs; p.o., per os; RV, residual volume; TLC, total lung capacity; 6MWD, 6min walk distance.

### Radiotracer production

[^11^C]NES and [^68^Ga]Ga-FAPI-46 were used as radioligands to assess regional NE and FAP expression as measures of inflammation and remodeling activity, respectively. Synthesis of [^11^C]NES was performed as described previously^13^. A fully automated manufacturing of [^68^Ga]Ga-FAPI-46 was developed using a synthesis platform (Modular PharmLab, Eckert & Ziegler Eurotope, Berlin, Germany) with disposable cassette system (C4-GA-PEP). It included ^68^Ga elution from the pharmaceutical grade generator (GalliaPharm, Eckert & Ziegler Radiopharma GmbH), labelling synthesis, product purification, product formulation, sterile filtration via 0.22 µm filter, and sterile filter integrity test. The quality control regarding radiochemical purity (98.4±1.4%), chemical purity, and quantity (< 50 µg) was conducted using high performance liquid chromatography with UV- and radiodetectors connected in series. The reaction buffer consisted of acetate buffer, gentisic acid, and ascorbic acid. The product, [^68^Ga]Ga-FAPI46 was formulated in total volume of 7.09±0.08 mL containing saline, PBS, EtOH (< 5%), and ascorbic acid to suppress the radiolysis. [^15^O]Water was produced according to the standard procedure for clinical diagnostic use at the PET Centre. All tracer production was in accordance with Good Manufacturing Practices and follows the guidelines in the European Pharmacopoeia and the manufacturing license from Swedish Medical Products Agency.

### PET/CT imaging

PET/CT scans were performed 23–30 months after COVID-19 diagnosis.

PET/CT scans were obtained using a digital time-of-flight hybrid PET/CT system (Discovery MI-5; GE Healthcare) with a 25-cm axial field of view. During each scan, the subjects were positioned supine.

Initially, lung perfusion was evaluated. A low-dose CT scan was conducted for anatomical localization and attenuation correction. Patients received a controlled bolus injection of 400 MBq of [^15^O]water, which was administered as 5 mL at a rate of 1 mL/min, followed by 35 mL of saline at a rate of 2 mL/min. Simultaneously, a 4-minute dynamic chest PET scan was initiated, with the following frame sequence: 1 × 10, 8 × 5, 4 × 10, 2 × 15, 3 × 20, and 2 × 30 s. After at least 15 minutes, inflammation was assessed by [^11^C]NES PET scan. A dynamic chest PET scan lasting 25 minutes (with frames of 1 × 10, 8 × 5, 4 × 10, 2 × 15, 3 × 20, 4 × 30, 5 × 60, and 3 × 300 s) was initiated simultaneously with a controlled bolus injection of 5 MBq/kg of [^11^C]NES (mean injected dose: 4.73 ± 0.13, approximately 2-4µg/mL of ^11^C-NES). Following this dynamic scan, whole-body low-dose CT and static PET scans of [^11^C]NES (3 minutes per bed position) were acquired.

Finally, carbon-11 activity was allowed to decay, and three hours after the injection of [^11^C]NES (equivalent to 9 half-lives ofcarbon-11), patients were scanned to assess remodeling activity with [^68^Ga]Ga-FAPI-46. Similar to the [^11^C]NES PET scan, a dynamic chest PET scan lasting 50 minutes (with frames of 1 × 10, 8 × 5, 4 × 10, 2 × 15, 3 × 20, 4 × 30, 5 × 60, 4 × 300 and 2 × 600 s) was initiated simultaneously with a controlled bolus injection of 3 MBq/kg of [^68^Ga]Ga-FAPI-46 (mean injected dose: 2.80±0.68, corresponding to approximately 2-4µg of [^68^Ga]Ga-FAPI-46). Following this dynamic scan, whole-body low-dose CT and static PET scans of [^68^Ga]Ga-FAPI-46 (3 minutes per bed position) were acquired. Dynamic and static whole-body PET/CT images were reconstructed as previously described^9^. The [^11^C]NES production failed due to technical reasons for the patient 003, who thus was scanned only with [^68^Ga]Ga-FAPI-46.

### [^11^C]NES, [^68^Ga]Ga-FAPI-46 and [^15^O]water data analysis

PET/CT images for [^11^C]NES and [^68^Ga]Ga-FAPI-46 were analyzed using PMOD 4.0 software (PMOD Technologies). For [15O]water aQuant Research software (Medtrace Pharma A/S) was used, as described previously^9^. Volumes of interest (VOIs) were delineated in the lung CT images. All the signals observed in the lungs on the PET images, with a mean standardized uptake value (SUV_mean_) scale ranging from 0.75 to 2 on the final frame of the PET scans (1200-1500s and 2400-3000s for [^11^C]NES and [^68^Ga]Ga-FAPI-46 respectively), were identified as VOIs for inflammation (I.V.) or remodeling volume (R.V.). The SUV lower threshold of 0.75 was chosen because it allowed for the subtraction of background uptake in the lungs of all patients. During the scans, the subjects are in a supine position, which can lead to dependent atelectasis accumulating along the basis posterior and a gravitational effect on the distribution of radiotracers in this same area. As a result, this part of the lung was delineated alone, and it was excluded from the I.V. or R.V. The signal observed around the lobar arteries and veins was also excluded from the I.V. or R.V.

In the lung, a VOI was drawn in the anterior part to represent the background uptake. The whole lung was also delineated (comprising the totality of the lungs, including atelectasis).

VOIs of other organs of interest (aorta, spleen, liver, bone marrow, bone, and muscle) were also delineated, and the results from this analysis are presented in supplementary data.

Similarly, VOIs were delineated on the whole-body PET/CT images, and the data are presented in the supplementary results.

Representative PET/CT images with VOIs are provided in the supplementary data to illustrate the method used (Fig.S1).

The results from the [^11^C]NES and [^68^Ga]Ga-FAPI-46 analyses are presented as follow: SUV_mean_ (mean SUV of voxels inside the VOI), SUV_max_ (SUV on the highest image pixel in the VOI), SUV_median_ (median SUV of voxels inside the VOI), and SUV_tot_ (SUV_mean_ multiplied by the VOI volume).

### Proximity Extension analysis (PEA)

Blood samples were obtained from healthy controls (n=2), patients with severe COVID-19 (n=4), and patients with long-COVID-19 (n=6). The samples were collected in EDTA tubes before any radiotracer injection on the day of the PET scans. The blood samples from healthy controls and patients with severe COVID-19 was taken during an earlier study evaluating [^11^C]NES binding in lung of these cohorts.^9^ Plasma was isolated by centrifuging blood in EDTA tubes at 4500 rpm for 5 minutes at 4°C. The plasma was extracted into cryotubes and stored at −80°C until it was used for Olink^®^ Target 96 analysis.

The frozen plasma samples were thawed on ice. 25 µL of plasma was deposited at the bottom of each V-shaped well in a 96-well plate. Each sample from healthy controls and long-COVID-19 patients was measured in four replicates, while the severe-COVID-19 samples were measured in triplicate. Plasma protein concentrations were analyzed using the Olink^®^ Target 96 inflammation panel by the SciLifeLab Unit of Affinity Proteomics in Uppsala, Sweden. Only samples that passed the quality control tests were reported. The results were expressed as normalized protein expression (NPX) on a log2 scale.

### Statistical analysis

The demographic and clinical characteristics of patients with long-COVID-19 are summarized in Table 1, presenting the median and the percentiles 25 and 75, for continuous variables and frequencies (expressed as the number of patients and percentages) for categorical variables. Between-group variations in plasma protein expression and comparison of tracer uptake and clinical parameters in a high and low uptake group were assessed using Kruskal-Wallis and uncorrected Dunn’s post-hoc tests. The correlation between PET-derived measurements, pulmonary function tests, 6 minutes walking tests (6MWTs), clinical information, and Olink^®^ proteomics measurements was analyzed using Spearman’s rank correlation. Z-scores were calculated using NPX and then plotted on a heatmap. Similar proteins and samples were clustered on the heatmap generated as suggested by the Olink**^®^** statistical analysis application.

Due to the small size of the study population, the p-values obtained from the statistical tests were considered exploratory. A significance level of 0.05 was used for all comparisons. Statistical analyses were conducted using GraphPad Prism 9.3.1 software (GraphPad Software, San Diego, CA, USA).

## Results

### Patients’ demographics

Six male patients (age 64 ± 7 [median ± range] years) who experienced persistent shortness of breath and reduced diffusion capacity for carbon monoxide (DLCO <75%) 20–30 months after severe COVID-19 illness were enrolled. Two patients were diagnosed with chronic kidney disease (CKD), and two diagnosed with type 2 diabetes mellitus (T2D). One patient did not perform the 6WMT.

### Multi-tracer PET/CT scans in long-COVID-19 patients’ lungs

PET scan results using [^11^C]NES and [^68^Ga]Ga-FAPI-46 showed variability among patients, ranging from minimal to pronounced accumulation of the tracer. The dynamic PET scans revealed that for both tracers, there was a peak uptake in the lungs from 0s to 40s or 50s (for [^68^Ga]Ga-FAPI-46 and [^11^C]NES, respectively) before reaching a stable uptake, following a similar dynamic uptake pattern as seen in the aorta (Fig. S2). The median I.V. represented 25.8% (IQR 12.9-30.9%) of the whole lung volume, while the median R.V. was 18.8% (IQR 8.9-33.89%) (Table 2). The SUV_mean_ in these volumes derived from the last frame of the dynamic scan, showed a median of 1.17 (IQR 1.14-1.37) in the I.V. and 1.04 (IQR 0.95-1.58) in the R.V.. Figure 1A illustrates a patient with high tracer uptake and another with low lung tracer uptake, which is representative of the results obtained. Patients 001, 002 and 003 showed low uptake, while patients 004, 005, and 006 exhibited high lung uptake (Fig. S3). For instance, patient 006 exhibited higher SUV_tot_ in both I.V. and R.V. than patient 001 (1029 and 765; 211 and 286, respectively). Lung perfusion, as measured with [^15^O]water, was not reduced in any of the scanned patients (perfusion scans are available in the supplemental data, Fig.S3). In areas exhibiting signs of reticulation, honeycombing, and ground glass opacity (GGO), we observed high remodeling activity with elevated levels of [^68^Ga]Ga-FAPI-46 and low levels of [^11^C]NES (Fig. 1B,C).

**Figure 1.**
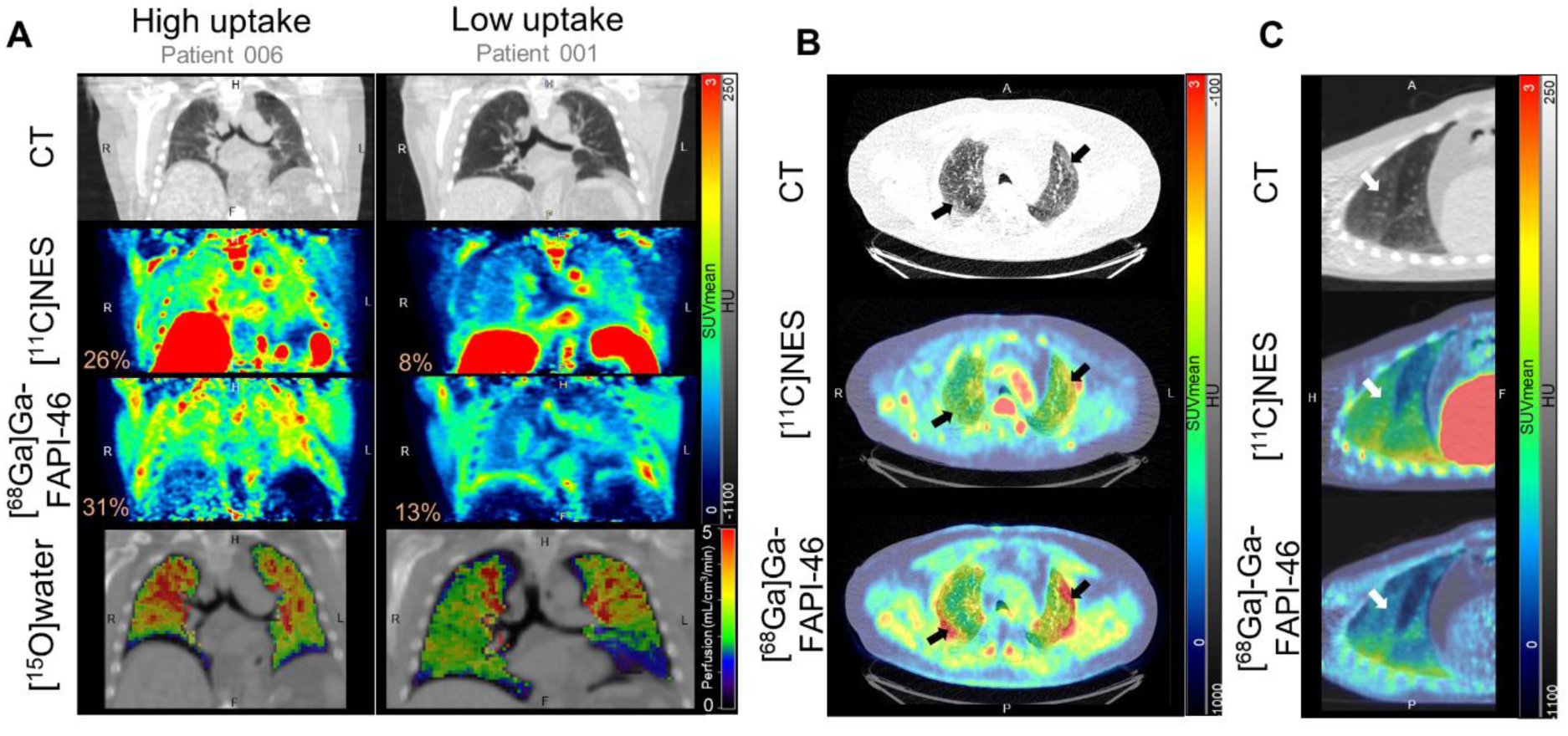
PET/CT scans in long-COVID-19 patients. PET images are from the last frame of the dynamic chest PET scans (1200-1500 s and 2400-3000 s for [^11^C]NES and [^68^Ga]Ga-FAPI-46, respectively). (A) Difference in uptake between a patient (patient 006) with high lung uptake for both tracers and a patient (patient 001) with low lung uptake (coronal sections). The percentages represented in orange are the per cent of inflammation ([^11^C]NES) or remodeling ([^68^Ga]Ga-FAPI-46) volume in the total volume of lungs. (B) [^11^C]NES and [^68^Ga]Ga-FAPI-46 uptake in areas of high remodeling activity, as indicated by the black arrows (patient 006)(transverse sections). (C) [^11^C]NES and [^68^Ga]Ga-FAPI-46 uptake in ground-glass opacity areas as indicated by white arrows (patient 005)(sagittal sections).

**Table 2.**
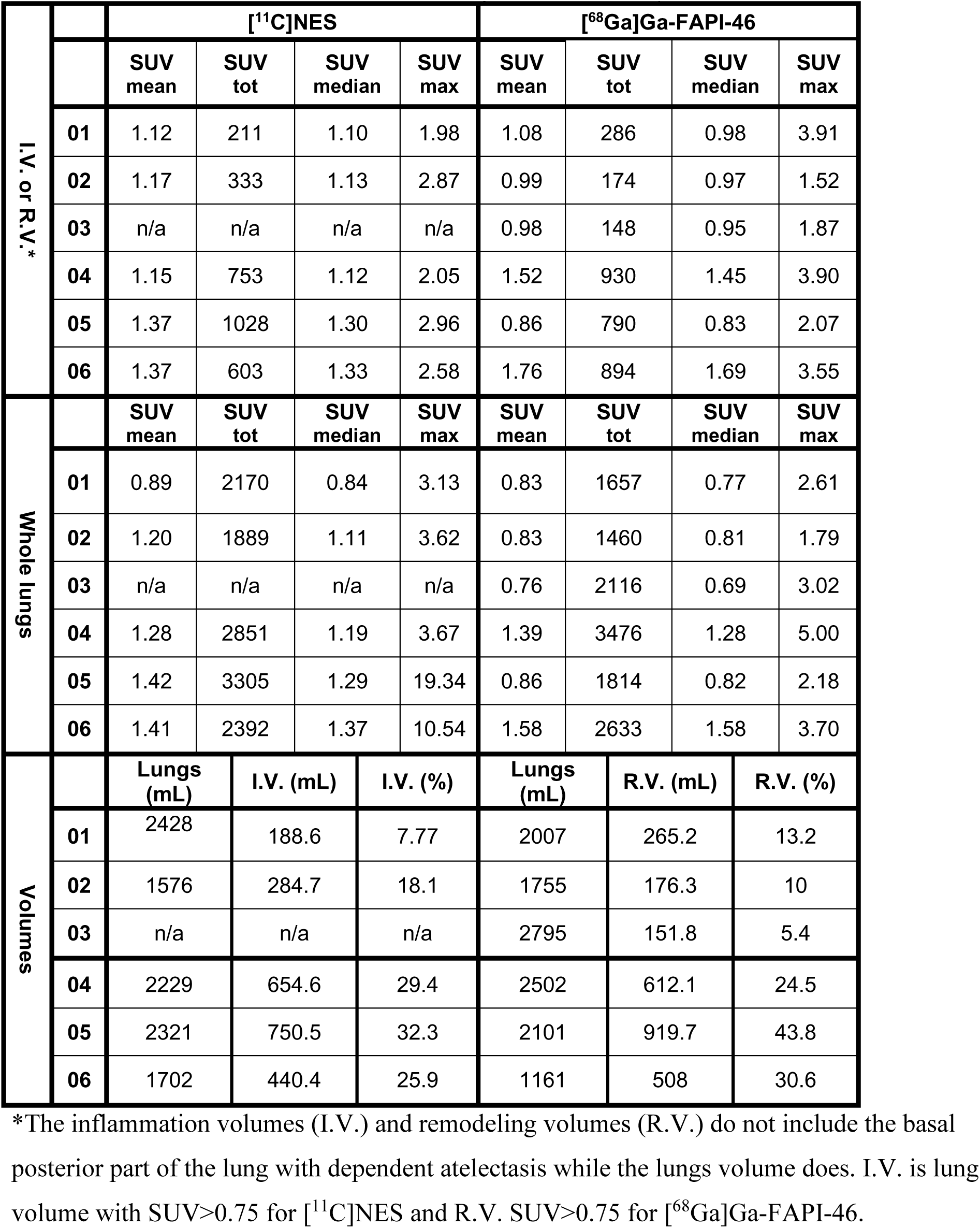
PET/CT derived measures in all long-COVID-19 subjects.

The tracers’ uptake in lungs and other organs for all patients are detailed in supplementary data (Table S1, Fig.S3). Overall, [^68^Ga]Ga-FAPI-46 shows lower uptake in most of the measured organs compared to [^11^C]NES. The [^11^C]NES exhibited significant uptake in n the liver and gallbladder, the measured radioactive in the [^11^C]NES scan represents radiolabeled metabolites. No radioactivity was seen in the kidney after about 30 min, but high levels in the urinary bladder representing excretion of intact tracer and metabolites.

Patient 004, diagnosed with type 2 diabetes mellitus T2D and CKD, exhibited nearly double [^68^Ga]Ga-FAPI-46 uptake in the pancreas compared to the other patients (SUV_mean_ of 2.42 and average SUV_mean_ of 1.19±0.13, respectively) (Table S1, Fig.S5). Additionally, the [^11^C]NES uptake in pancreas of the same patient was the second highest after patient 005, who had the highest NES uptake. Patient 003, who also had T2D, showed an average uptake of [^68^Ga]Ga-FAPI-46 and [^11^C]NES in the pancreas, unlike Patient 004. Patient 004’s scans also showed a higher uptake of both tracers in the left kidney compared to the average uptake in kidneys of patient without kidney disease (Fig.S5). Patient 002 with CKD did not show higher kidney uptake compared to patients without a CKD diagnosis.

Both tracers exhibited significant uptake in the right maxillary sinus of patient 006, who was diagnosed with sinusitis (Fig. S6).

### Proximity Extension analysis (PEA) of peripheral inflammation markers

Proteomic profiling of plasma from healthy patients, severe-COVID-19 patients, and long-COVID patients using the Olink**^®^** inflammation 96 panel allowed us to observe heterogeneous inflammation profiles between the groups (Fig.2). Among the 19 proteins with an NPX difference above 0.8 between the healthy and long-COVID-19 samples, only two proteins were higher in the healthy control than in the long-COVID-19 samples: EN-RAGE (extracellular newly identified receptor for advanced glycation end-products binding protein, also named S100A12) and Interleukin-1 alpha (IL1α). IL1α was also reduced compared to controls in severe-COVID-19, while EN-RAGE was higher in both control and long-COVID-19 groups. Severe-COVID-19 protein expression tended to follow long-COVID-19 patterns for the cytokines: interferon gamma (IFN-γ), IL8, and IL1α and for the chemokines: monocyte chemoattractant protein (MCP)-1, MCP-2, MCP-4, C-C chemokine ligand (CCL)11, and CCL4. Some of these proteins, such as the chemokines C-X-C motif chemokine ligand-9 (CXCL9), CXCL10, MCP3, CCL3, CDCP1 (CUB domain containing protein 1), and EN-RAGE, tend to be higher in plasma from severe COVID-19 compared to long-COVID-19. In contrast, fibroblast growth factor 21 (FGF21), FGF5, IL5, and matrix metalloprotease 10 (MMP10) levels tended to be higher in the long-COVID-19 group than in the severe-COVID-19 group.

**Figure 2.**
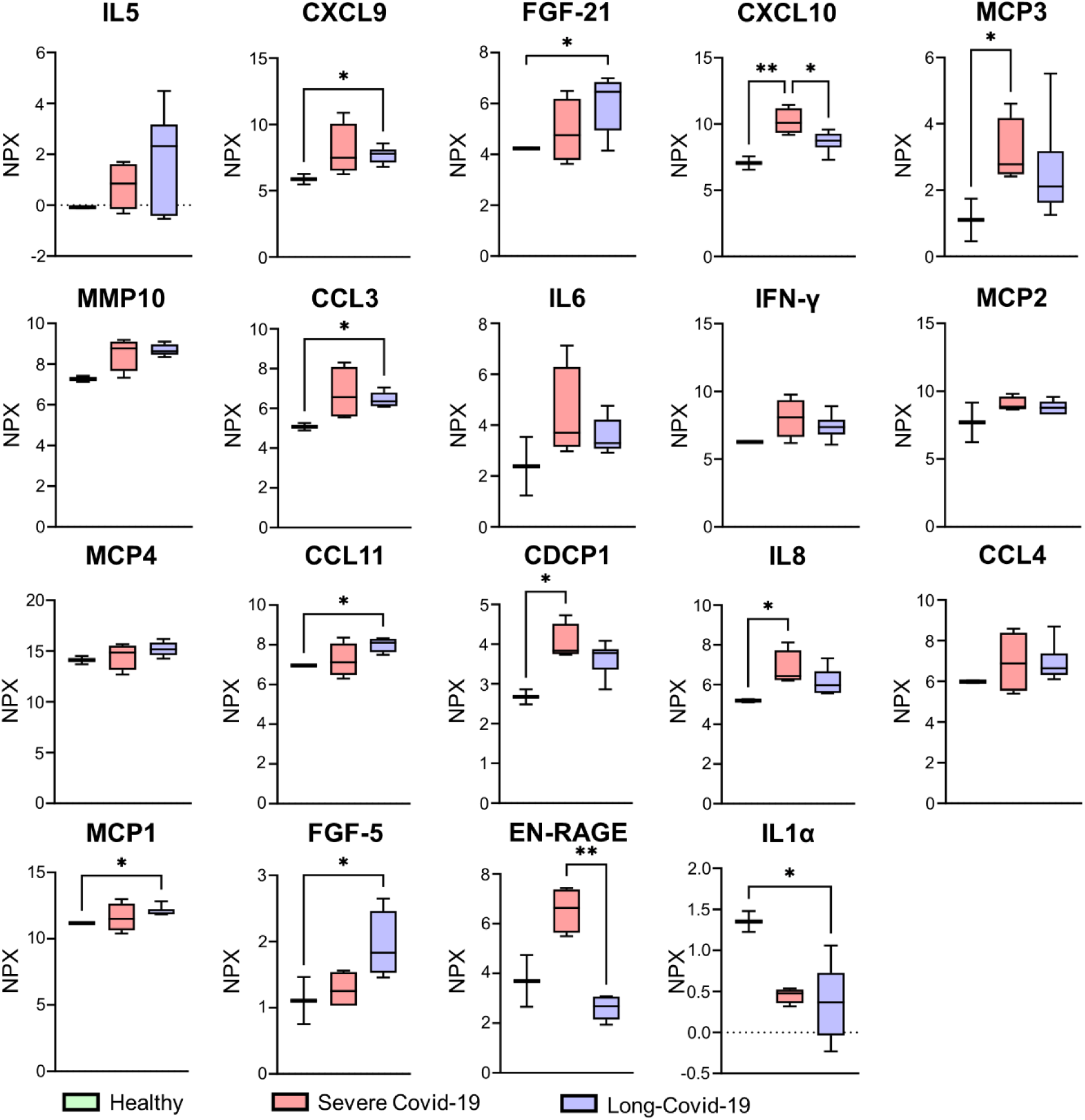
Proteomic profiling of plasma using PEA. Only the proteins that have an NPX difference > 0.8 between the healthy controls and long-COVID-19 patients are represented. Box-whisker plots showing differentially expressed inflammatory proteins in plasma from healthy controls (n=2). severe COVID-19 (n=4) and long-COVID-19 patients (n=6). Level of significance: * p<0.05; ** p<0.01 and. **** p<0.0001 (Kruskal-Wallis and uncorrected Dunn’s post-hoc tests).

### Correlations between the different parameters studied

The study also investigated the relationships between PET-derived measures, clinical parameters, and plasma proteomic profiles from the Olink® inflammation panel (Fig.3, S7, and S8). High uptake levels of [^11^C]NES were associated with high uptakes of [^68^Ga]Ga-FAPI-46. Clinical parameters such as duration of IV, intensive care unit (ICU) stay, and the 6MWD increased with the volume of tracers’ uptake of both tracers. No significant correlations were observed between the lung parameters and tracer uptake, except for the total lung capacity percentage (TLC%), which was associated with low tracer uptake, especially for [^11^C]NES ([^11^C]NES: SUV_tot_, r=-0.79, and SUV_mean_, r=-0.84).

**Figure 3.**
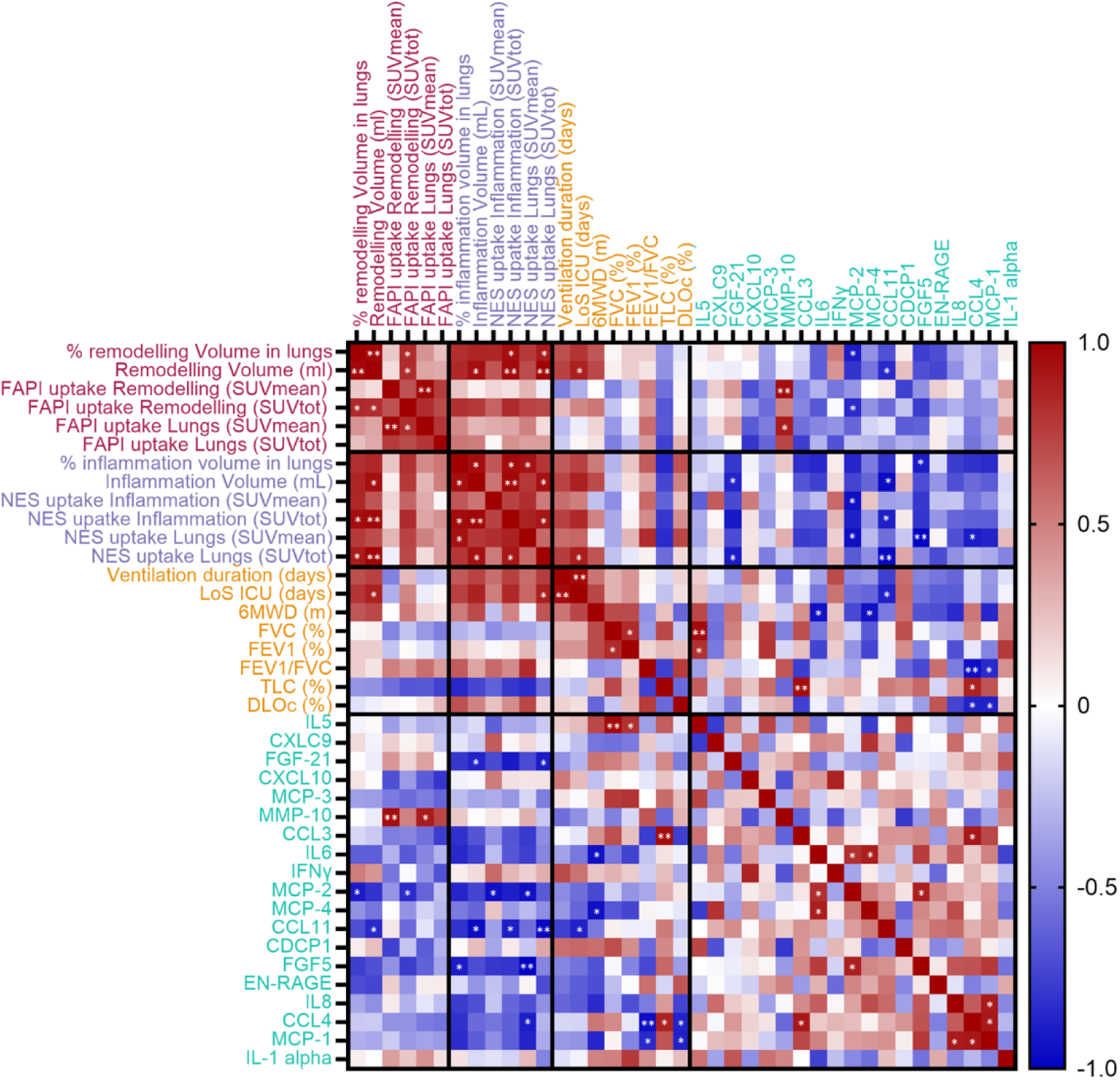
Correlation matrix between PET-derived measures. clinical parameters and plasma proteomic profile. Pearson correlation coefficients (*r*) were calculated for combinations of PET data, results of clinical lung function and lung plasma proteomic. Color intensity indicates the strength of the correlations. Level of significance: * p<0.05; and ** p<0.01.

The study also investigated the relationship between PET-derived measures and plasma proteomic analysis. For several inflammatory biomarkers, there was no apparent correlation between their plasma levels and radiotracer uptake, including IL6 and IL8.

Conversely, proteins, such as FGF5, CCL11, and MCP-2 tended to show decreased plasma levels associated with high SUVt_ot_ uptake. Notably, CCL11 showed a significant negative correlation with [^11^C]NES SUV_tot_ (r=-0.88). MCP-2 plasma levels were negatively correlated with SUV_tot_ for both tracers ([^11^C]NES: r=-0.88; [^68^Ga]Ga-FAPI-46: r=-0.83). FGF21 negatively correlated with [^11^C]NES SUV_tot_ (r=-0.90), but not with that of [^68^Ga]Ga-FAPI-46 (r=0.16).

## Discussion

In this pilot investigation, we focused on patients with reduced lung capacity as a consequence of long-COVID-19. Utilizing a triad of PET scans, we non-invasively monitored perfusion ([^15^O]water), inflammation ([^11^C]NES), and tissue remodeling ([^68^Ga]Ga-FAPI-46) in these patients. The results of the [^15^O]water PET scan showed that the respiratory symptoms linked to long-COVID-19 did not have an association with lung perfusion. All patients showed lung uptake of the inflammation radiotracer [^11^C]NES and the remodeling radiotracer [^68^Ga]Ga-FAPI-46, albeit at varying intensity levels. The group being studied showed a dichotomy in radiotracer uptake, enabling the categorization of individuals into two groups based on the low and high uptake of these radiotracers.

^68^Ga-labeled FAPI radiotracers have been extensively studied as an oncologic imaging tool due to the expression of FAP on the surface of cancer-associated fibroblasts^14,15^. Beyond its oncological applications, FAP also plays a role in tissue remodeling and is expressed on activated fibroblasts. Consequently, ^68^Ga-labeled FAPI radiotracers have been investigated for pathologies characterized by uncontrolled scarring, such as fibrosis. Specifically, investigations in the fibrotic lungs of patients with ILD have revealed that ^68^Ga-FAPI uptake is exclusively present in areas undergoing fibrotic tissue remodeling^16^. The use of ^68^Ga-labeled FAPI PET scans for COVID-19 lung sequelae has been limited, with only a few patients studied. Notably, one reported case described diffuse bilateral basal posterior lung uptake of [^68^Ga]Ga-FAPI-46 in a patient experiencing COVID-19 pneumonia sequelae^17^. However, it is important to consider that this region is also susceptible to dependent atelectasis when patients are scanned in a supine position. Another study involving [^68^Ga]Ga-FAPI-46 and six patients with long-COVID-19 reported a mean SUV_max_ of 2.75±0.32 in the suspected fibrotic area^18^. In the present study, the average SUV_max_ in the R.V. 55 min post-injection closely matched this observation (2.81±1.00, range [1.52;3.91]). Emphasizing the importance of considering both SUV_max_ and the location/volume of tracer uptake, our data suggests that a high SUV_max_ may not necessarily indicate a high R.V. or I.V. This volume uptake is associated with areas of interest, such as GGO or reticulation (Fig. 1B,C), whereas SUV_max_ can be affected by atelectasis within the VOI (Table 2).

In contrast to ^68^Ga-labeled FAPI radiotracers, the [^11^C]NES radiotracer was newly described in an inflammation mouse model^13^ and an acute respiratory distress syndrome (ARSD) pig model^19^ before being investigated in a first-in-man study involving patients with severe COVID-19^9^. This study found that the uptake of [^11^C]NES was higher in areas showing GGO and consolidations than in healthy lung tissue. This finding is consistent with our observations in long-COVID-19 patients, which showed similar SUV_mean_ in intravenous or opaque lung tissue (Fig. 1B, C). In lung areas with decreased air content, perfusion is normally decreased due to hypoxic vasoconstriction. However, unlike in the previous study, our evaluation of lung perfusion using [15O]water did not show a clear reduction in GGO and consolidation in the lungs of patients with long COVID-19 (Fig.1 and S3). This could contribute to hypoxia and dyspnea

Expanding on prior research, we further compared tracer uptake with plasma proteomics and clinical parameters.

Clinical parameters used to evaluate lung function, such as TLC and FVC, tended to decrease with increasing tracers’ uptake, while the ventilation duration and the LoS ICU increased. These measurements seemed to have a negative correlation with tracer uptakes and could potentially predict the results of PET scans using [^11^C]NES and [^68^Ga]Ga-FAPI-46. However, research on larger cohort is needed to validate these observations.

The proteomics analysis indicated increased systemic inflammation in the severe cases of COVID-19, with elevated plasma levels of EN-RAGE, IL-6, MCP-3, IFN-γ, CXCL10, and CXCL9 compared to the levels found in healthy control plasma, as supported by the literature^20,21^. However, the immunology of long COVID-19 in plasma seems more complex. Plasma levels of cytokines such as IL-8, IL-6, and IFN-γ were higher than those observed in healthy controls, consistent with previous findings^8,22^. However, among individuals with long-COVID-19, these plasma biomarkers levels were not associated to lung tracer uptakes, suggesting discrepancy between systemic and local inflammatory processes. Pro-inflammatory chemokines driving neutrophil recruitment, CCL8 (MCP-2), tended to be inversely associated with tracer uptake, including [^11^C]NES. The plasma levels of inflammatory markers might be influenced by various pathologies. For example, a high level of FGF-21 appears to be associated with low [^11^C]NES SUV_tot_. However, it has been reported that the serum level of this biomarker is increased in individuals with obesity and T2D^23^. While the plasma inflammatory profile seems to vary between long-COVID-19 and healthy patients, it does not seem to predict the uptake of NES and FAPI in the lungs of long-COVID-19 patients.

In addition to lung uptake, significant tracer uptake was observed in other organs with pathology, including kidney and pancreas of a patient (Patient 004) with T2D and CKD (Fig.S5, Table.S1). Yet, while two patients studied were diagnosed with this T2D (patients 003 and 004), only one patient showed increased uptake of tracers. This could be related to the different treatment received as patient 004 is treated with glucagon-like peptide-1 (GLP-1) agonist in addition to Sodium-dependent glucose cotransporter (SGLT) antagonist. Some studies associated GLP-1 agonists with pancreatitis^24,25^, but this observation is still debated.^26^ FAP is involved in T2D pathogenesis and is considered a therapeutic target of interest in T2D because its enzymatic activity deactivates FGF21, which acts as a protective metabolic regulator^27–29^. FAPI radiotracer uptake was recently described in the pancreas of a patient with fulminant type 1 diabetes.^30^ Furthermore, Patient 004, who also suffers from CKD, showed a significant uptake in his left kidney. [^68^Ga]Ga-FAPI-04 PET scanning, conducted on a small group of patients with kidney fibrosis, revealed elevated uptake in the kidneys compared to that in healthy patients^31^, consistent with findings from preclinical studies^32^.

Other sites with inflammatory activity such as in the maxillary sinus of a patient with sinusitis showed an elevated uptake of both tracers (Fig.S6). In addition, compared with [^68^Ga]Ga-FAPI-46, significant uptake of [^11^C]NES was observed in the liver, and gallbladder, indicating hepatobiliary excretion. Additionally, a high uptake was observed in the spleen and bone marrow, where resident neutrophils are produced (Table S1, Fig. S4). These observations correlate with findings from preclinical and clinical studies using this tracer^9,13^. The uptake observed in the stomach could also be explained by the presence of neutrophils.

The main limitation of this single-center pilot study is the small sample size, as only six patients were included, which means that the statistical tests conducted are purely exploratory. Given the limited cohort size and the lack of diversity, as only male patients were studied, further research with larger and more diverse cohort is required to validate and extend these observations.

The pathophysiological mechanisms responsible for long-COVID-19 remain unknown, and clear guidelines for its treatment are lacking. In addition, diagnosing pulmonary fibrosis based on CT findings remains a challenge. There is a risk of either over-diagnosing lung fibrosis on follow-up CT scans or missing it altogether because CT features of lung fibrosis are not always clearly present^33,34^. Invasive histological confirmation is usually not feasible, as biopsy rarely is indicated and can associated with severe side effects. Therefore, there is an urgent need for non-invasive clinical diagnostic tools to assess the inflammation and remodeling status in the lungs of patients with long-COVID-19.

In this context, the PET scans described in this study using the radiotracers [^11^C]NES and [^68^Ga]Ga-FAPI-46 have significant potential to address this challenge. These scans could play a crucial role in stratifying patients for the selection of therapeutic strategies and identifying individuals who may benefit from anti-fibrotic or anti-inflammatory treatments. The use of PET scans may provide valuable insights for a more precise and targeted approach to managing patients with long-COVID-19.

## Conclusion

In conclusion, this single-center first-in-humans study demonstrated that lung perfusion, neutrophil-mediated inflammation and tissue remodeling can be assessed in patient with long-COVID-19 using PET imaging. These findings contribute to a more comprehensive understanding of the complex pathophysiology of long-COVID-19 and offer insights that may guide therapeutic decisions, particularly in addressing individual variations within the patient population.

## Take home points

### Study Question

Can PET triad scans using [^15^O]water, [^11^C]NES, and [^68^Ga]Ga-FAPI-46 assess lung perfusion, neutrophil activity, and remodeling activity in the lungs of long-COVID-19 patients, and do these scan results correlate with clinical parameters and inflammatory proteomics?

### Results

Lung perfusion remained unaffected, but clear evidence of neutrophil and remodeling activity was observed in areas with suspected fibrosis. The results from the PET scans could not be efficiently associated with clinical parameters or inflammatory proteomics.

### Interpretation

This pilot study provides encouraging data on [^11^C]NES and [^68^Ga]Ga-FAPI-46 PET scans for assessing lung inflammatory and remodeling status in patients with lung capacity sequelae in the context of long-COVID-19. These findings contribute to a better understanding of the biological processes underlying long-COVID-19.

## Financial/nonfinancial disclosures

Olof Eriksson is an employee and a minority shareholder of Antaros Tracer AB. Otherwise, the authors have nothing to disclose.

## Acknowledgments

The authors would like to acknowledge the staff at the Uppsala University Hospital PET center for technical excellence. Sofie Bioscience provided the precursor FAPI-46, under a Material Transfer Agreement.

## Financial support

This work has been supported by the Swedish Heart-Lung Foundation (grant No. 20210012). O.W.s and O.E.s positions are funded by the Swedish Research Council (2020-02312) and Science for Life Laboratory.

## Data availability

The datasets generated during and/or analyzed during the current study are available from the corresponding author upon reasonable request.

## Abreviations

ARSD: acute respiratory distress syndrome
BMI: body mass index
CCL: C-C chemokine ligand
CDCP1: CUB domain containing protein 1
CKD: chronic kidney disease
CXCL: C-X-C motif chemokine ligand
COPD: chronic obstructive pulmonary disease
COVID-19: coronavirus disease 2019
CT: computed tomography
DLCO: diffusing capacity for carbon monoxide
EN-RAGE: extracellular newly identified receptor for advanced glycation end-products binding protein
FAP: fibroblast activation protein
FAPI: fibroblast activation protein inhibitor
FEV1: forced expiratory volume
FEV1/FVC: Tiffeneau-Pinelli index
FGF: fibroblast growth factor
FVC: forced vital capacity
GGO: ground glass opacity
GLP-1: glucagon-like peptide-1
ICU: intensive care unit
IFN-γ: interferon gamma
IL: interleukin
ILD: interstitial lung disease
IV: invasive ventilation
I.V.: inflammation volume
LoS: length of stay
MCP: monocyte-chemotactic protein
MMP: matrix metalloproteinase
NE: neutrophil elastase
NES: GW457427
NET: neutrophil extracellular traps
NPX: normalized protein expression
NSAID: nonsteroidal anti-inflammatory drugs
PET: positron emission tomography
RV: residual volume
R.V.: remodeling volume
SARS-Cov2: severe acute respiratory syndrome coronavirus 2
SUV: standardized uptake value
TLC: total lung capacity
T2D: type 2 diabetes
VOI: volume of interest
6MWD: 6 minutes walking distance
6MWT: 6 minutes walking test

## Notes

### Author Declarations

The Swedish Ethical Review Authority gave ethical approval for this work (Dnr.: 2021-03218).

