## Supplementary data for "Exploring impacts of long-COVID-19 on the lungs: A triad of PET scans for perfusion, inflammation and tissue remodeling"

**Short title:** Multi-parametric PET scans in long-COVID-19

Olivia Wegrzyniak[1], Olof Eriksson[1], Emil Ekbom[2], Robert Frithiof[2], Michael Hultström[2,3], Mark Lubberink[4], Jonathan Sigfridsson[4], Irina Velikyan[1,5], Viola Wilson[1,5], Gunnar Antoni[5,6]*, Miklos Lipcsey[1,6]*

### Table of contents

Fig.S1……………………………………………………………………………….2

Fig.S2……………………………………………………………………………..3

Fig.S3……………………………………………………………………………..4

Fig.S4……………………………………………………………………………..5

Fig.S5……………………………………………………………………………..6

Fig.S6……………………………………………………………………………..7

Fig.S7……………………………………………………………………………..8

Fig.S8……………………………………………………………………………..9

Table S1……………………………………………………………..……………..10

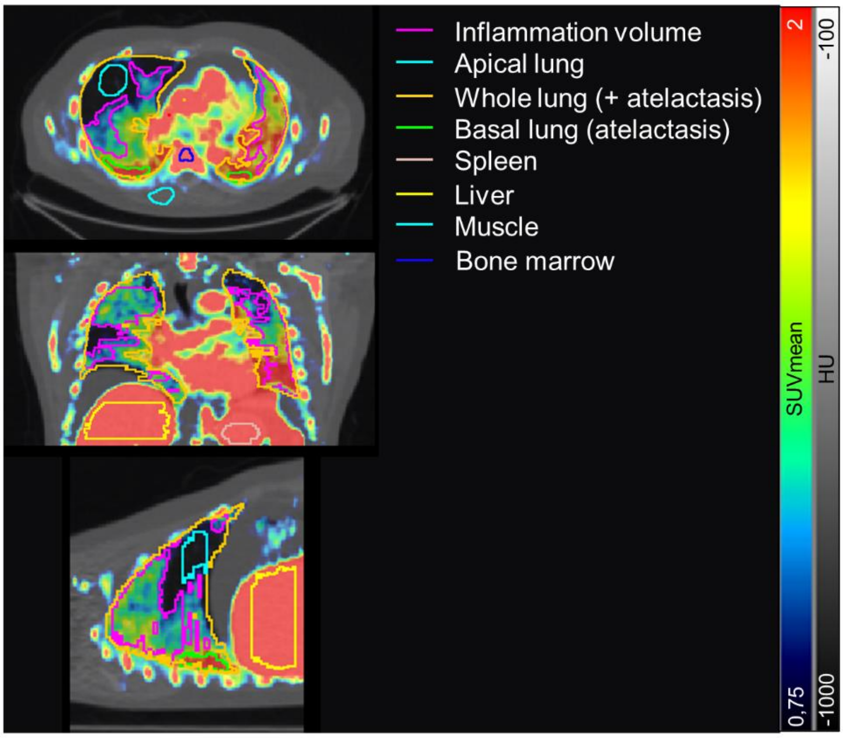


**Figure S1.** Illustration of the methods used to draw the VOIs of the lungs in patient 005. The SUV scale was from 0,75 to remove the background signal in the lung. Atelectasis (drawn here as "basal lung" in green) and uptake from lobar arteries and veins removed from the inflammation volume VOI.

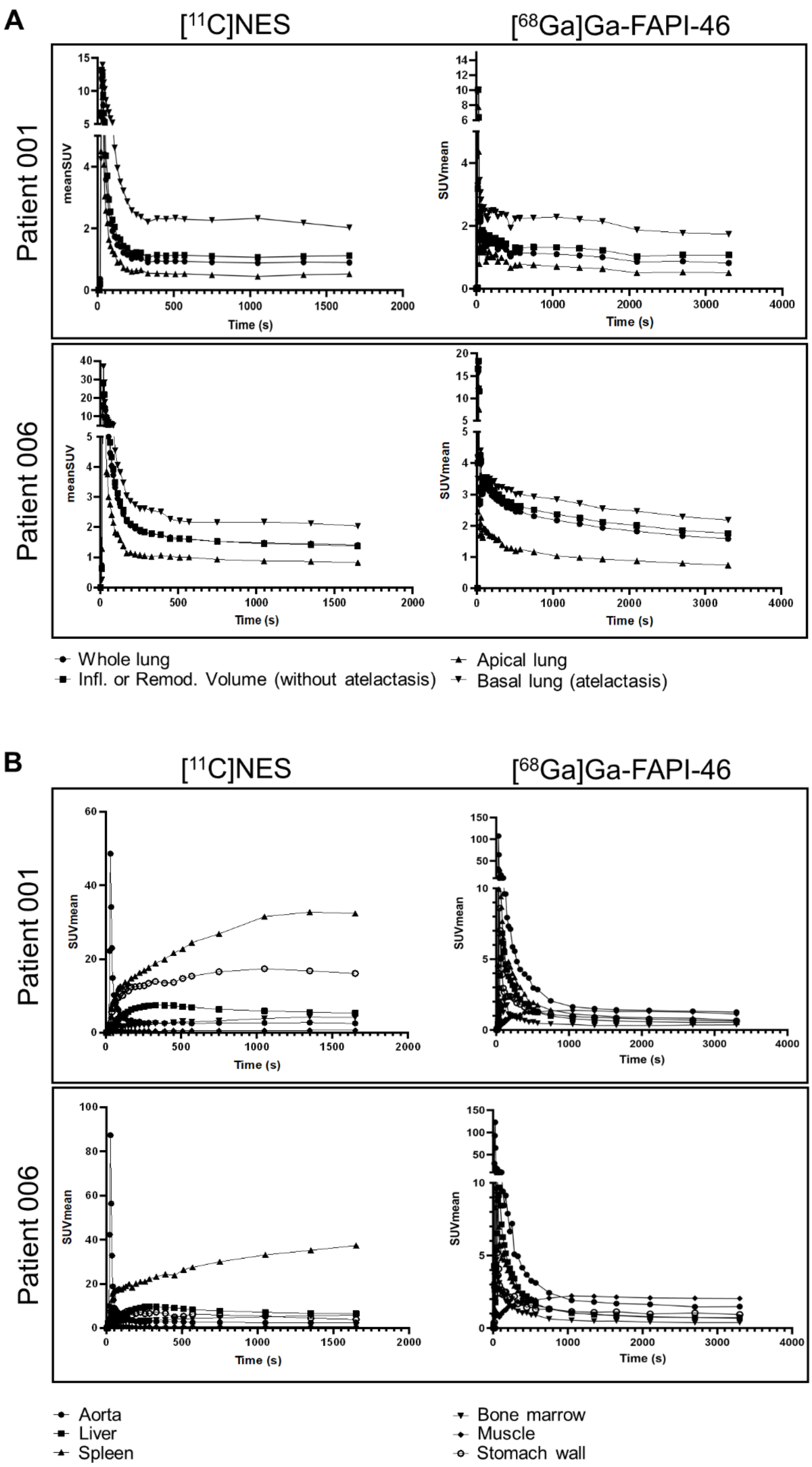


**Figure S2.** Time activity curves of [$^{11}$C]NES and [$^{68}$Ga]Ga-FAPI-46 uptake in lungs (A) and other organs (B).

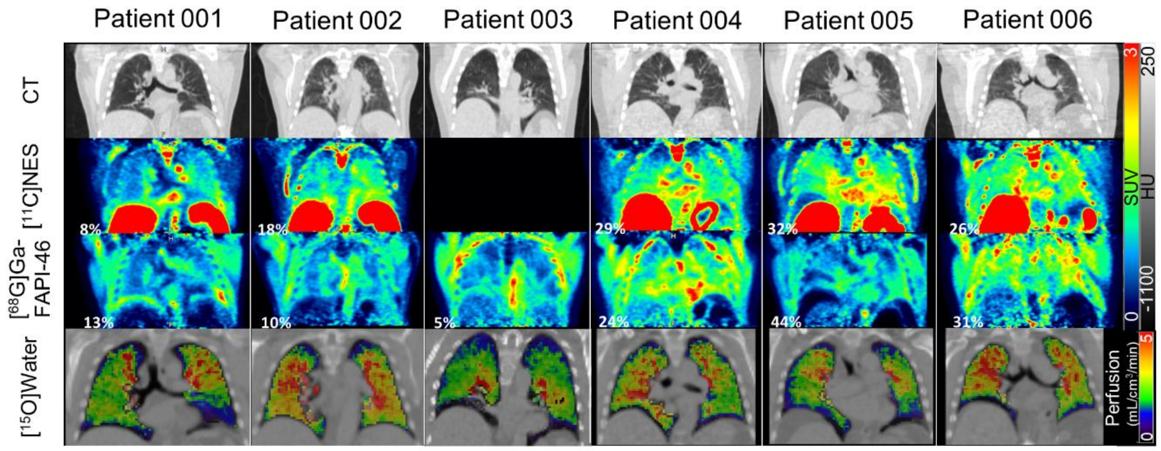


**Figure S3.** PET/CT images of [$^{11}$C]NES, [$^{68}$Ga]Ga-FAPI-46 and [$^{15}$O]water uptake in all patients. PET images are from the last frame of the dynamic chest PET scans (1200-1500 s and 2400-3000 s for [$^{11}$C]NES and [$^{68}$Ga]Ga-FAPI-46 Respectively).

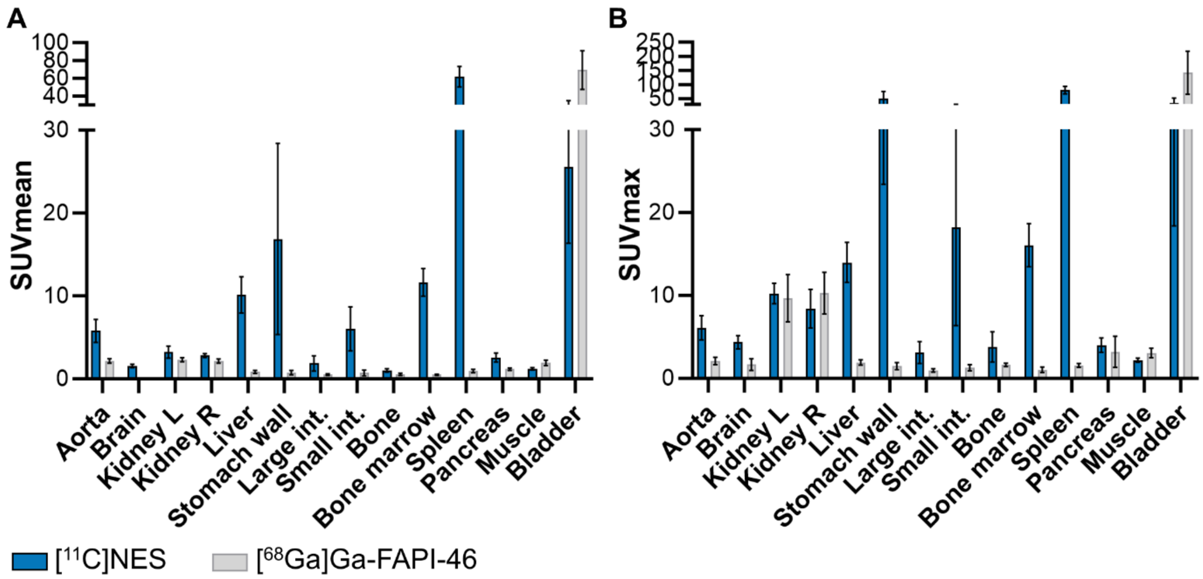


**Figure S4.** Mean $SUV_{mean}$ (A) and $SUV_{max}$ (B) of [$^{11}$C]NES and [$^{68}$Ga]Ga-FAPI-46 derived from static full body scans.

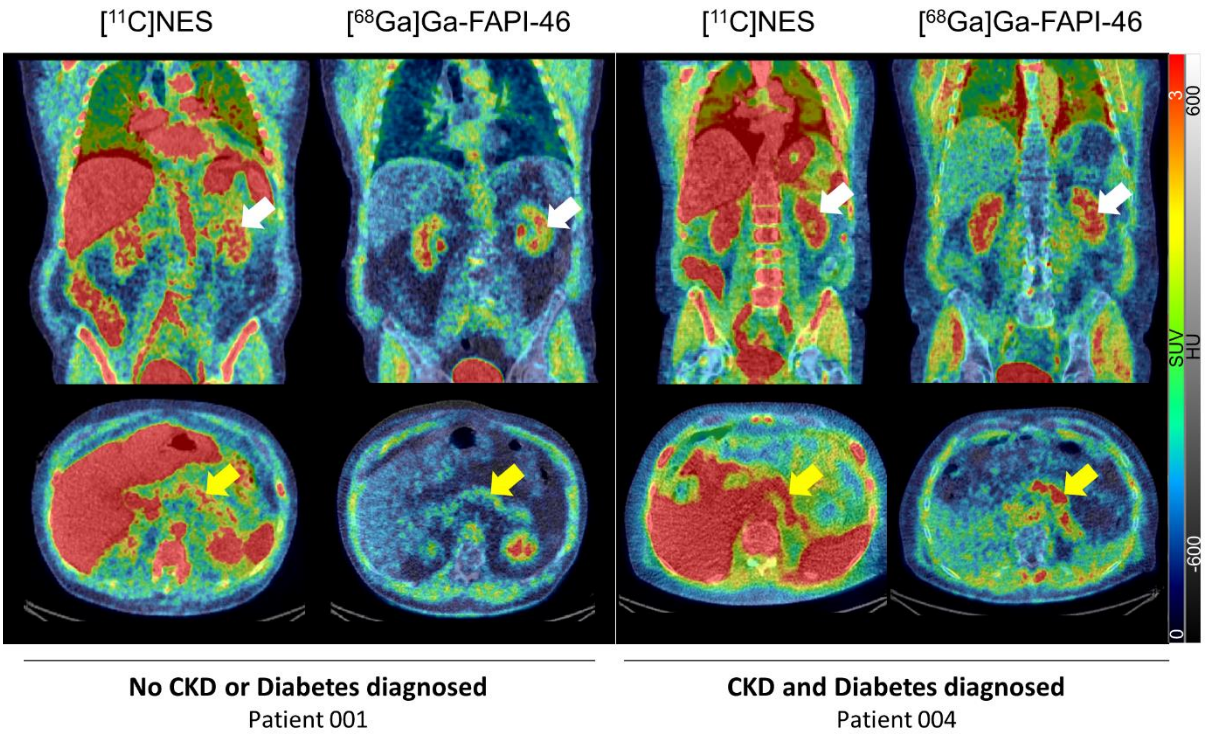


**Figure S5.** PET/CT images of [$^{11}$C]NES and [$^{68}$Ga]Ga-FAPI-46 uptake in kidneys and pancreas of a patient non-diagnosed for CKD and diabetes (left) and one who was diagnosed with these diseases (right). White arrows indicate a kidney, and yellow arrows the pancreas.

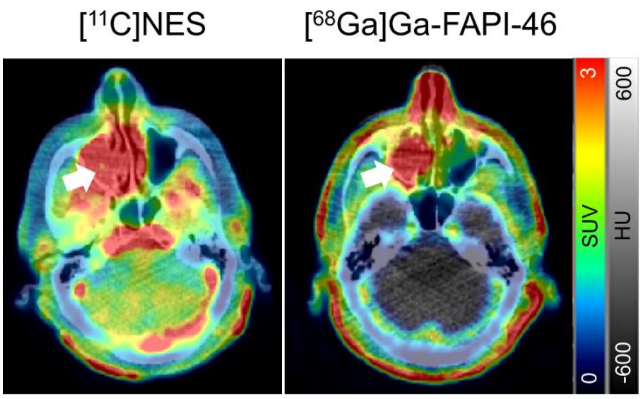


**Figure S6.** PET/CT images of [$^{11}$C]NES and [$^{68}$Ga]Ga-FAPI-46 uptake in the maxillary sinus of a patient with sinusitis. White arrows indicate the significant uptakes in the right maxillary sinus.

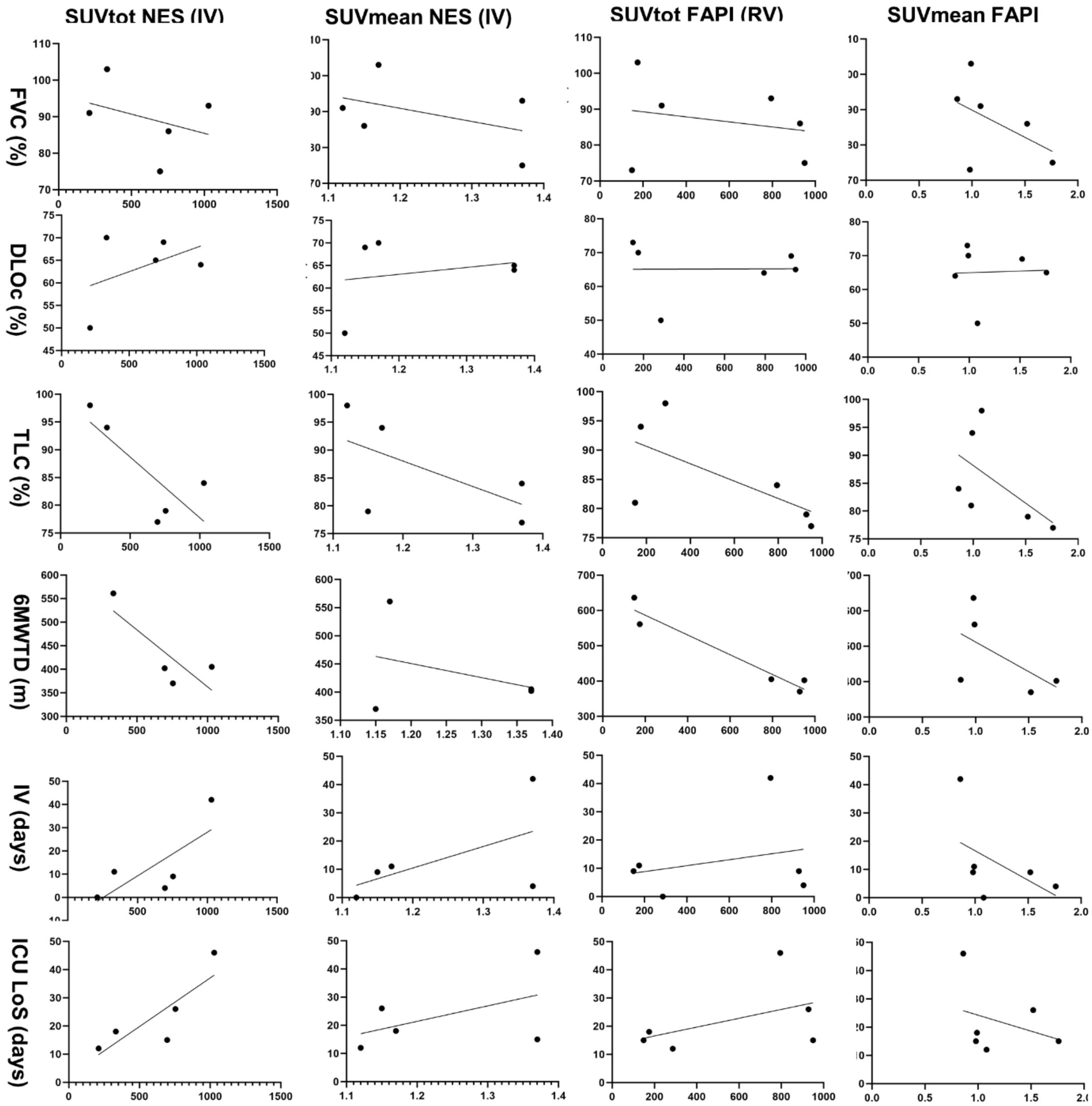


**Figure S7.** Scatter plots with liner regression of PET derived measured compared to clinical parameters.

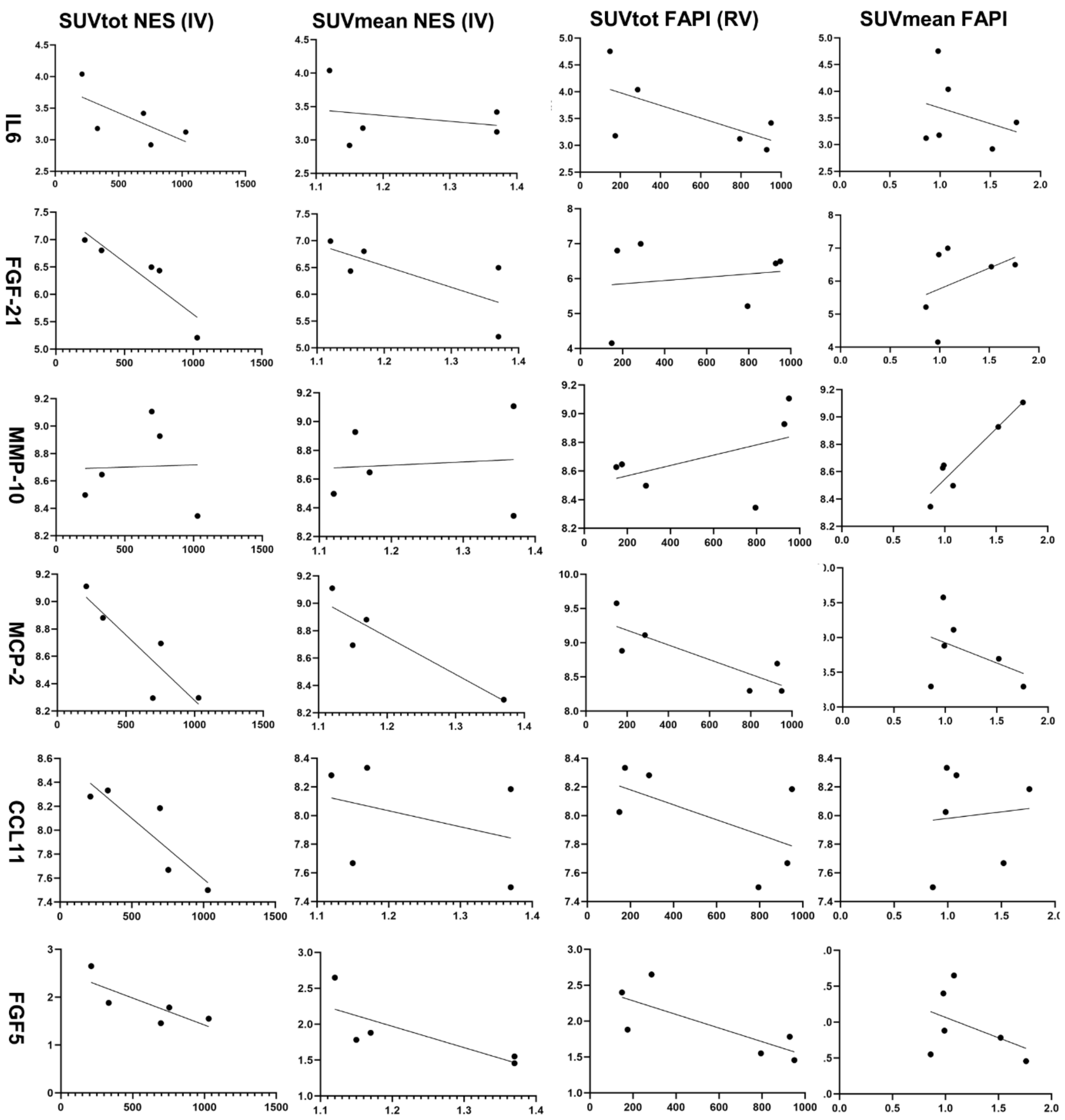


**Figure S8.** Scatter plots with liner regression of PET derived measured compared to plasma level of inflammatory biomarkers from Olink® inflammation 96 panel.

**Table S1:** Static full body PET/CT derived SUV measures in all long-COVID-19 subjects.

| | | Patient 001 | Patient 002 | Patient 003 | Patient 004 | Patient 005 | Patient 006 |
|---|---|---|---|---|---|---|---|
| **[$^{11}$C]NES** | | | | | | | |
| **Brain** | | 1.51 | 1.30 | n/a | 1.65 | 1.55 | 1.87 |
| **Aorta** | | 7.67 | 3.96 | n/a | 6.43 | 5.66 | 5.21 |
| **Kidney** | **Left** | 3.00 | 2.40** | n/a | 4.05** | 3.58 | 2,68 |
| | **Right** | 2.95 | 2.65** | n/a | 2.95** | 3.87 | 2.55 |
| **Liver** | | 8.81 | 7.65 | n/a | 12.40 | 12.41 | 9.36 |
| **Spleen** | | 76.88 | 49.62 | n/a | 61.34 | 69.58 | 51.79 |
| **Pancreas** | | 2.23 | 2.25 | n/a | 2.96* | 3.36 | 2.50 |
| **Stomach wall** | | 35.34 | 9.92 | n/a | 10.64 | 21.66 | 7.22 |
| **Large intestine** | | 2.97 | 2.73 | n/a | 1.13 | 1.56 | 1.04 |
| **Small intestine** | | 4.92 | 2.67 | n/a | 6.64 | 9.96 | 6.02 |
| **Bone** | | 0.89 | 1.33 | n/a | 1.15 | 0.86 | 0.98 |
| **Bone marrow** | | 10.30 | 13.91 | n/a | 12.71 | 11.21 | 10.00 |
| **Bladder** | | 22.58 | 16.32 | n/a | 23.55 | 24.35 | 41.05 |
| **Muscle** | | 1.09 | 1.13 | n/a | 1.27 | 1.13 | 1.46 |
| **[$^{68}$Ga]GaFAPI-46** | | | | | | | |
| **Brain** | | 0.02 | 0.02 | 0.01 | 0.02 | 0.02 | 0.03 |
| **Aorta** | | 2.55 | 1.97 | 2.08 | 2.34 | 1.83 | 2.30 |
| **Kidney** | **Left** | 2.09 | 2.08** | 2.20 | 2.79** | 2.64 | 2.27 |
| | **Right** | 1.93 | 2.16** | 2.07 | 2.38** | 2.51 | 2.17 |
| **Liver** | | 0.69 | 0.66 | 0.93 | 1.16 | 0.72 | 0.94 |
| **Spleen** | | 0.83 | 1.12 | 0.79 | 1.32 | 0.92 | 0.86 |
| **Pancreas** | | 1.05 | 1.10 | 1.26* | 2.42* | 1.24 | 1.39 |
| **Stomach wall** | | 0.86 | 0.55 | 0.75 | 1.03 | 0.32 | 1.03 |
| **Large intestine** | | 0.50 | 0.67 | 0.49 | 0.53 | 0.36 | 0.63 |
| **Small intestine** | | 0.28 | 0.59 | 0.90 | 1.21 | 0.48 | 0.82 |
| **Bone** | | 0.44 | 0.64 | 0.77 | 0.53 | 0.54 | 0.43 |
| **Bone marrow** | | 0.47 | 0.49 | 0.45 | 0.69 | 0.49 | 0.48 |
| **Bladder** | | 98.49 | 43.81 | 79.06 | 65.50 | 45.72 | 83.64 |
| **Muscle** | | 1.36 | 1.83 | 2.30 | 2.34 | 1.87 | 2.03 |

*Patients diagnosed with type 2 diabetes, **patients diagnosed with chronic kidney diseases.
